# eYoung men’s experiences of violence and poverty and the relationship to sexually transmissible HIV: a cross sectional study from rural South Africa

**DOI:** 10.1101/2024.09.08.24313251

**Authors:** Andrew Gibbs, Esnat Chirwa, Guy Harling, Natsayi Chimbindi, Jaco Dreyer, Carina Herbst, Nonhlanhla Okesola, Osee Behuhuma, Nondumiso Mthiyane, Kathy Baisley, Thembelihle Zuma, Theresa Smit, Nuala McGrath, Lorraine Sherr, Janet Seeley, Maryam Shahmanesh

**Author notes:** Correspondence: Department of Psychology, Washington Singer Building, Perry Road, Exeter, UK, EX4 4QG. Meetings analysis presented Poster presentation: AIDS2025, Munich, Germany, July 2024.

## Abstract

**Background:** Young (ages 18-35 years) men are inadequately engaged in HIV prevention and treatment globally, including in South Africa, increasing the likelihood of them having sexually transmissible HIV (i.e. living with HIV but with high viral loads). We sought to understand how men’s experiences of poverty and violence, impacted on transmissible HIV, directly or indirectly via mental health and substance misuse.

**Setting:** Rural communities in northern KwaZulu-Natal, South Africa.

**Methods:** Cross-sectional population-based random selection (September 2018-June 2019), assessing transmissible HIV (living with HIV and viral load ≥400 copies/mL) via dried blood spots, and socio-demographic data. Structural equation models (SEM), assessed direct and indirect pathways from food insecurity and violence experience to transmissible-HIV, with mediators common mental disorders, alcohol use, gender inequitable attitudes and perceptions of life chances.

**Results:** 2,086 young (ages 18-36 years) men and 8.6%(n=178) men had transmissible HIV. In SEM no direct pathways between food insecurity, or violence experience, and transmissible HIV. Poor mental health and alcohol use mediated the relationship between violence experience and food insecurity and transmissible HIV. Life chances also mediated the food insecurity to transmissible HIV pathway.

**Conclusions:** There was a high level of transmissible HIV in a representative sample of young men. The analysis highlights the need to address both the proximate ‘drivers’ poor mental health and substance misuse, as well as the social contexts shaping these among young men, namely poverty and violence experience. Building holistic interventions that adequately engage these multiple challenges is critical for improving HIV among young men.

## Introduction

Men are inadequately engaged in HIV prevention and treatment in generalised HIV epidemics ^1-4^, negatively impacting their health and the health of their sexual partners ^5^. The steps leading towards viral suppression – HIV testing, access to treatment and treatment adherence – are often referred to as the HIV treatment cascade ^6^. In generalised HIV epidemics men are lost more often than women at each of these steps ^1,4,5,7^. This pattern of low engagement in treatment is seen in South Africa where young men living with HIV (aged 25-35 years) have the poorest viral suppression rates (66.3%) of all groups nationally ^8^. These rates are even lower in some settings, for example among young men (aged 16-29 years) in a trial of Treatment as Prevention in rural KwaZulu-Natal, less than 80% knew their HIV status, 25% were on treatment, and only 18% virally suppressed ^9,10^.

Improving the health of men and their partners requires a focus on those at greatest risk of HIV transmission and acquisition. An important emerging definition is “sexually transmissible HIV”, a composite measure capturing both recent (incident) HIV and those who have not (successfully) achieved viral suppression; both groups are more likely to transmit HIV sexually because of high viral load ^11,12^. This definition resonates with the central role of antiretrovirals in achieving good health and reducing or eliminating onward transmission of HIV ^12^. There are, however, three broad categories of reasons why sexually transmissible HIV is inadequately addressed among men in generalized epidemic settings.

First, health systems are frequently unresponsive to men. Material barriers make it challenging for men (and women) to access health services, e.g. poverty and the cost of transport to clinics undermine access ^1^. Food insecurity further impacts people’s ability to take HIV treatment ^13^. Additionally, when men work this may conflict with access to healthcare because clinics have limited opening hours or long waiting times ^14^. Moreover, health services often actively exclude men as they are targeted at women and children ^2^.

Second, dominant forms of gender inequitable masculinity undermine men’s engagement throughout the cascade ^1^. This includes reducing willingness to test for HIV because of concerns about how an HIV positive diagnosis could undermine men’s social identities as strong, self-reliant and a provider if testing positive ^15-17^. Other studies suggest men may struggle to take medication regularly, as it challenges notions of their well-self ^14,16^.

Third, men’s poor mental health and substance misuse can increase their prevalence of sexually transmissible HIV ^18-20^. Poor mental health can reduce linkage to care post-testing ^21^. Moreover, poor mental health and substance use can also impact on men’s ability to regularly take medication, increasing their likelihood of having a detectable viral load ^22^. Additionally, research has suggested that a lack of hope and future orientation may further impact on people living with HIV’s engagement in HIV care and treatment ^23^.

Importantly, men’s poor mental health and substance misuse needs to be located within the social contexts in which men live, particularly poverty, violence and men’s masculinities. Research has demonstrated that poverty, particularly food insecurity, is strongly associated with poor mental health ^24^ and substance use ^25^. Similarly, research has described how men’s own experiences of violence are associated with poorer mental health and substance misuse ^26^. Finally, in contexts of poverty and violence, men are more likely to develop less gender equitable masculinities ^27^, which may include greater alcohol use, less gender equitable attitudes and likely to draw on ideas of strength and resist accessing health care.

In this paper we draw on a population-based sample of young men living in a rural setting with high HIV prevalence and incidence to understand the factors associated with transmissible HIV. We hypothesise that among this population sexually transmissible HIV is likely shaped by the broad social context of men’s experiences of violence and poverty, but that these social factors will also be mediated by men’s mental health, substance use and gender attitudes.

## Methods

### Setting

The study was conducted in the Hlabisa sub-district of uMkhanyakude district, in northern KwaZulu-Natal, South Africa. The area – the site of the long-running Africa Health Research Institute (AHRI) Health and Demographic Surveillance System ^28^ – is primarily rural with one large town. There is relatively little infrastructure or industry and high rates of unemployment. HIV incidence is high among young men in this area, with low uptake of HIV prevention services, HIV testing and HIV treatment ^9,10,29^.

### Design

This is a secondary analysis of a cohort of young men aged between 13 and 35 years at baseline, selected using a random stratified sample from the AHRI census of household residents in 2017 to create a population-representative sample. Baseline interviews were conducted in 2018 and 2019, with fieldwork teams identifying participants before conducting informed consent and face-to-face interviews. Data were collected in a pre-programmed REDCap ^30^ system in isiZulu.

### Measures

HIV serostatus at baseline was assessed through Dried Blood Spots (DBS). We initially categorised participants into three groups: not living with HIV; living with HIV but no detectable viral load (<400 copies/mL); and sexually transmissible HIV, which was defined as living with HIV and having a detectable viral load of□≥□400 copies/mL ^12^. In the main analysis we combined ‘not living with HIV’ and ‘living with HIV without a detectable viral load’ into transmissible HIV, compared to sexually transmissible HIV.

To understand men’s experience of past-year violence we used a 15-item scale, modified from the WHO Violence Against Women scale ^31^. We assessed men’s past year experience of emotional (3 items), physical (8 items) and sexual (4 items) violence, with the preparator being ‘anyone’ (Supplementary File: Table 1). Each item had a “yes or no” response. We treated each form of violence men could have experienced (i.e. emotional, physical, sexual) separately, whereby a positive response to one or more items in that group, was considered indicative of experiencing that form of violence. We also created a breadth score of violence experienced, through summing all items, and higher scores indicative of experiencing more forms of violence, following prior coding approaches ^32^.

**Table 1.**
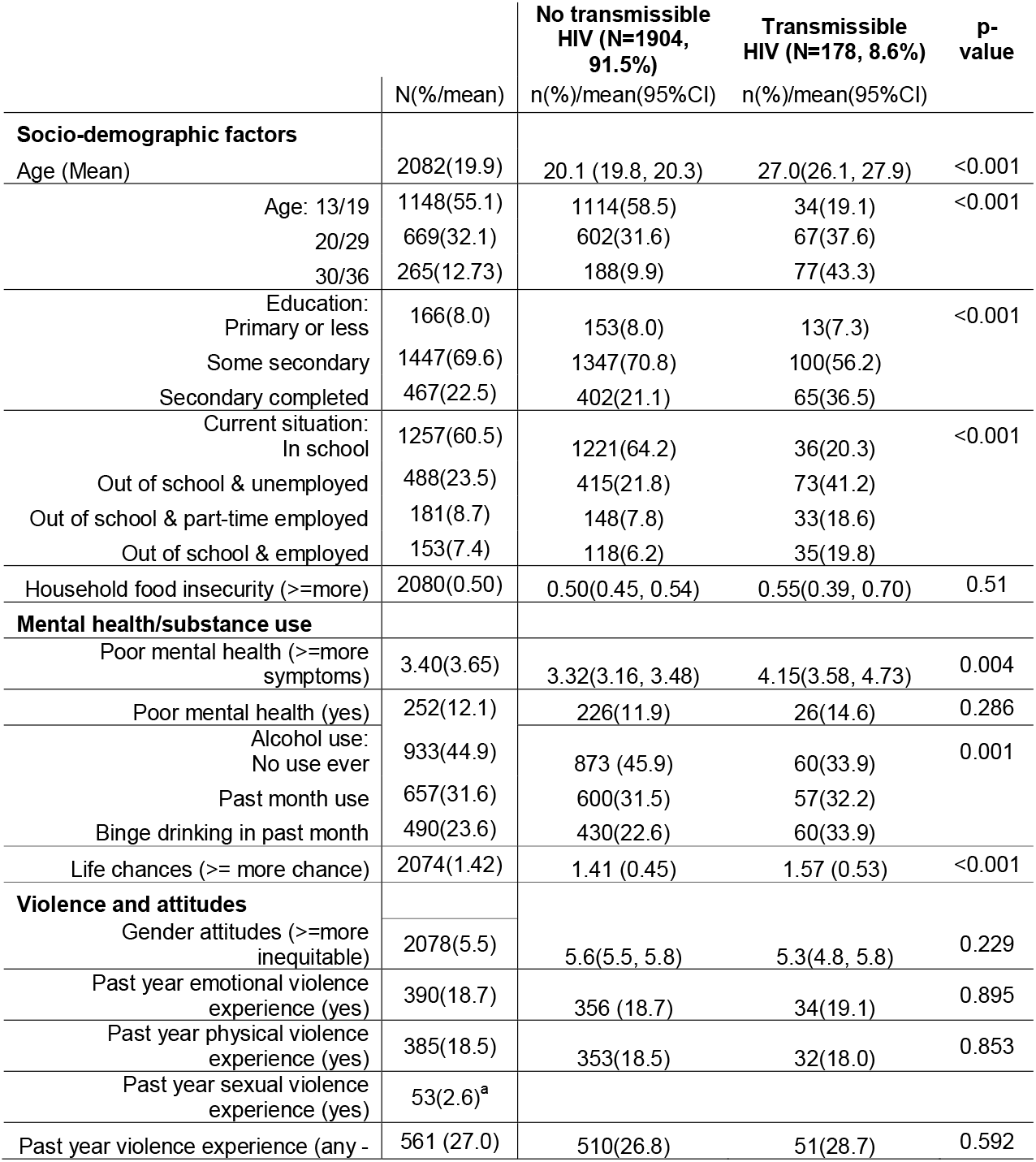
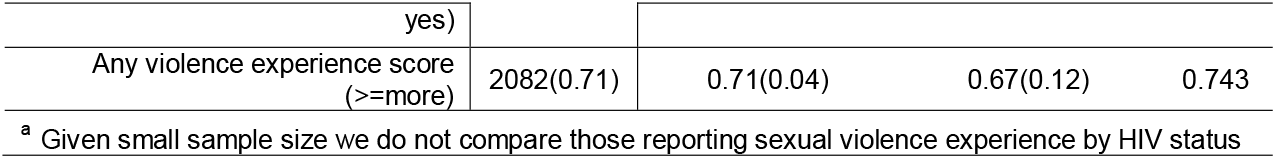
Sample description and descriptive association comparing those with transmissible HIV to those without transmissible HIV.

|  | N(%/mean) | No transmissible HIV (N=1904, 91.5%)<br>n(%)/mean(95%CI) | Transmissible HIV (N=178, 8.6%)<br>n(%)/mean(95%CI) | p-value |
| --- | --- | --- | --- | --- |
| **Socio-demographic factors** |  |  |  |  |
| Age (Mean) | 2082(19.9) | 20.1 (19.8, 20.3) | 27.0(26.1, 27.9) | <0.001 |
| Age: 13/19 | 1148(55.1) | 1114(58.5) | 34(19.1) | <0.001 |
| 20/29 | 669(32.1) | 602(31.6) | 67(37.6) |  |
| 30/36 | 265(12.73) | 188(9.9) | 77(43.3) |  |
| Education: |  |  |  |  |
| Primary or less | 166(8.0) | 153(8.0) | 13(7.3) | <0.001 |
| Some secondary | 1447(69.6) | 1347(70.8) | 100(56.2) |  |
| Secondary completed | 467(22.5) | 402(21.1) | 65(36.5) |  |
| Current situation: |  |  |  |  |
| In school | 1257(60.5) | 1221(64.2) | 36(20.3) | <0.001 |
| Out of school & unemployed | 488(23.5) | 415(21.8) | 73(41.2) |  |
| Out of school & part-time employed | 181(8.7) | 148(7.8) | 33(18.6) |  |
| Out of school & employed | 153(7.4) | 118(6.2) | 35(19.8) |  |
| Household food insecurity (≥more) | 2080(0.50) | 0.50(0.45, 0.54) | 0.55(0.39, 0.70) | 0.51 |
| **Mental health/substance use** |  |  |  |  |
| Poor mental health (≥more symptoms) | 3.40(3.65) | 3.32(3.16, 3.48) | 4.15(3.58, 4.73) | 0.004 |
| Poor mental health (yes) | 252(12.1) | 226(11.9) | 26(14.6) | 0.286 |
| Alcohol use: |  |  |  |  |
| No use ever | 933(44.9) | 873 (45.9) | 60(33.9) | 0.001 |
| Past month use | 657(31.6) | 600(31.5) | 57(32.2) |  |
| Binge drinking in past month | 490(23.6) | 430(22.6) | 60(33.9) |  |
| Life chances (≥ more chance) | 2074(1.42) | 1.41 (0.45) | 1.57 (0.53) | <0.001 |
| **Violence and attitudes** |  |  |  |  |
| Gender attitudes (≥more inequitable) | 2078(5.5) | 5.6(5.5, 5.8) | 5.3(4.8, 5.8) | 0.229 |
| Past year emotional violence experience (yes) | 390(18.7) | 356 (18.7) | 34(19.1) | 0.895 |
| Past year physical violence experience (yes) | 385(18.5) | 353(18.5) | 32(18.0) | 0.853 |
| Past year sexual violence experience (yes) | 53(2.6)<sup>a</sup> |  |  |  |
| Past year violence experience (any - | 561 (27.0) | 510(26.8) | 51(28.7) | 0.592 |
| yes) |  |  |  |  |
| Any violence experience score<br>(>=more) | 2082(0.71) | 0.71(0.04) | 0.67(0.12) | 0.743 |

<sup>a</sup> Given small sample size we do not compare those reporting sexual violence experience by HIV status

Poverty was assessed through the proxy measure of household food insecurity. First, people were asked if in the past year they, or someone in their household, had skipped a meal because of a lack of food. For those who answered ‘yes’, we then asked about frequency, with response categories, ‘every month’, ‘some months’ or ‘once or twice’. We then created a score to indicate food insecurity combining both items (range 0-3), with higher scores indicating greater insecurity, following an approach used in this population previously ^33^.

To understand mental health and substance use we asked three sets of questions. We used the 14-item Shona Symptom Questionnaire (SSQ-14) to assess general mental health ^34^.

The scale assesses past week symptoms of mental health via binary questions such as “Were you frightened by trivial things?”. The items are summed to create a total score, with higher scores indicating worse mental health. We used the standard cut-point of 8 or more items to be indicative of poor mental health ^34^.

We assessed perceptions of future life chances through 12 items drawn from a prior study in Kenya ^35^. Questions asked participants to assess the likelihood they would complete key milestones in life, such as ‘finish secondary school’, ‘have a happy family life’ and ‘be respected in the community’. Responses were ‘high, or already achieved’, ‘50:50’ or ‘low’. We summed items, and higher scores indicated a greater sense of future life chances.

Alcohol use was assessed with two items. First, respondents were asked whether they had drunk alcohol in the past 12 months. Among those reporting alcohol consumption, we asked how many days in the past month they had engaged in binge drinking (five or more drinks in one sitting). We recoded alcohol use as: never in the past 12 months; alcohol use but not binge drinking in the last month; and, binge drinking in the last month.

Gender attitudes were assessed using the Gender Equitable Men’s scale (GEMS) ^36^. Men were asked 30 items about a range of gender-related attitudes and practices, with responses options of “agree” or “disagree”. We conducted a principal component factor analysis, identifying many items with poor loading, finalising the scale using 17-items which cohered as a scale. Three broad groups of items were included, around sex (e.g., “It is the man who decides what type of sex to have”), children (e.g., “Only when a woman has a child is she a real woman”) and violence (e.g., “If a woman cheats on a man, it is okay for him to hit her”). Items were summed, and higher scores were indicative of more gender inequitable attitudes.

We assessed sociodemographic information, including age (as a continuous variable), highest completed or current (if still in school) education level (primary or less; secondary not completed; secondary completed) and current life situation (in school; out-of-school and unemployed; out-of-school and employed part-time; out-of-school and employed full time).

### Analysis

Descriptive analyses were conducted in STATA 18. We present summary statistics for the full sample, using numbers and percentages or means and 95% confidence intervals (95%CI) as appropriate. Our main analysis compares men without transmissible HIV (whether because HIV-negative or HIV-positive but with undetectable viral load) to those with transmissible HIV. We do this first, by descriptively assessing the differences between those without transmissible HIV and those with transmissible HIV (using t-tests or chi-squared tests and reporting p-values). Second, we then conducted path modelling in R using lavaan ^37^. To do this we first identified the theoretical relationship between variables assuming their directionality. Second, we tested each of these paths through regression within the Structural Equation Modelling (SEM) framework, and if regressions were significant, included these in the next step. Third, we fitted the full model of significant paths, and ran the model, and iteratively removed paths which were non-significant (p>0.05). We then tested Goodness of Fit, using Tucker-Lewis Index (TLI), Comparative Fit Index (CFI) and Root Mean Square Error of Approximation (RMSEA). We treated the outcome (transmissible HIV) as a binary variable. We initially included age as a continuous co-variate in the SEM, but the model would not converge, and as such age was excluded.

We conducted a series of supplementary analyses. First, we descriptively looked at the sample comparing those without HIV, those with non-transmissible HIV, and those with transmissible HIV. Second, we ran additional SEMs with different outcomes (people without HIV versus those with HIV, and HIV-negative compared to those with transmissible HIV).

### Ethics

The study received ethical approval from the Biomedical Research Ethics Committee at the University of KwaZulu-Natal and from the University College London’s ethics committee. The study was also approved by AHRI’s community advisory board. All participants aged 18 and above provided written informed consent to participate in the study. For those aged under 18, signed parental consent was obtained and signed assent from the participant.

## Results

In total 3,577 men were sampled and sought for interview, of whom 69.5% (n=2,487) were found and consented to participate. Those who refused participation were older (Supplementary File: Table 2). Among those who consented, biological HIV information was collected from 83.7% (n=2082). Those without HIV DBS data were older, more likely to have completed secondary education, have more gender equitable attitudes and less likely to have experienced any form of violence (Supplementary File: Table 3).

**Table 2.**
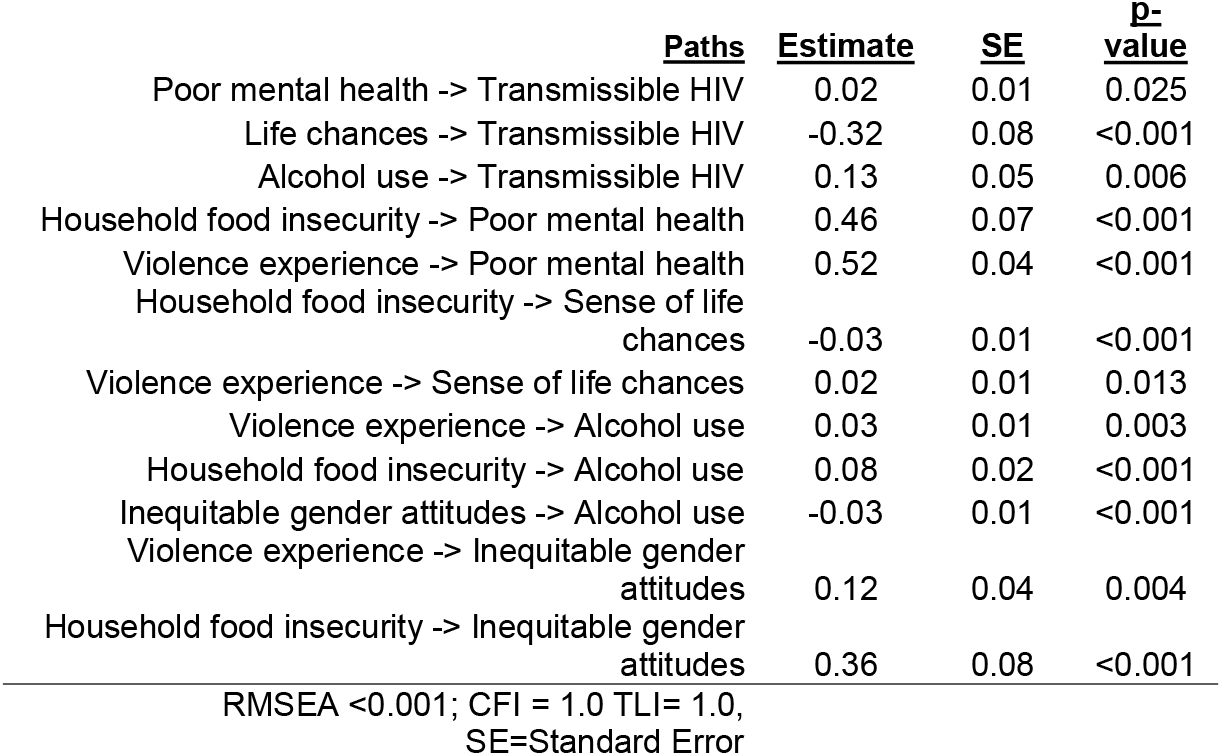
Structural relationships for transmissible HIV.

Among the 2082 male participants who provided baseline data and an HIV test (Table 1), 11.0% (n=229) were living with HIV. Among those HIV-positive, 78%(n=178) had transmissible HIV (i.e. viral loads >= 400) and 22% (n=51) did not have detectable viral loads. In the full analytic sample included, mean age was 21.2 years (range 13-36 years), and two-thirds (69.6%) reported having some secondary education. Three-fifths (60.5%) of respondents were still in school, a quarter (23.5%) were unemployed and the remaining were out of school and had some form of employment.

One in ten (12.1%) had poor mental health, and mean scores on this scale were: 3.40(SD=3.65, range 0-14). A quarter (23.6%) reported binge drinking in the past month, and almost half (44.9%) reported never drinking alcohol in the past year. Past year experience of violence was common: overall a quarter (27.0%) reported any form of violence, a fifth (18.7%) reported experiencing past year emotional violence, a fifth (18.5%) past year physical violence, and 1 in 50 (2.6%) past year sexual violence (2.0%, n=41 reported experiencing all forms of violence).

Table 1 compares those without transmissible HIV (91.5%) to those with transmissible HIV (8.6%). Compared to those without transmissible HIV, mean ages for those with transmissible HIV were higher, and a greater proportion were out of school, and reported binge drinking in the past month. Additionally, mean scores were higher for symptoms of poor mental health, and sense of lack of life chances among those with transmissible HIV compared to those without. Supplementary analyses (Supplementary File: Table 4) comparing those without HIV, to those with HIV with non-detectable viral loads, had higher mean ages, less likely to be in school, reported binge drinking in the past month and less gender inequitable attitudes. While those with transmissible HIV, were older, more likely to be out of school, reported binge drinking in the past month, less future chances and poorer mental health.

In the path model (Figure 1, Table 2) there were no direct pathways between recent violence experience, nor food insecurity and transmissible HIV. Food insecurity to transmissible HIV was mediated via mental health, life chances and alcohol use, whereby greater household food insecurity was associated with increases alcohol use, poorer mental health and reduced life chances, and these in turn were associated with an increased likelihood of sexually transmissible HIV. Experience of past year violence was similarly mediated to sexually transmissible HIV, via mental health and alcohol use, whereby more violence experiences were associated with poorer mental health and more alcohol use, and then sexually transmissible HIV. In contrast to expectations, recent violence experience was associated with a higher sense of life chances (i.e. those who had experienced more violence, reported better life chances).

**Figure 1.**
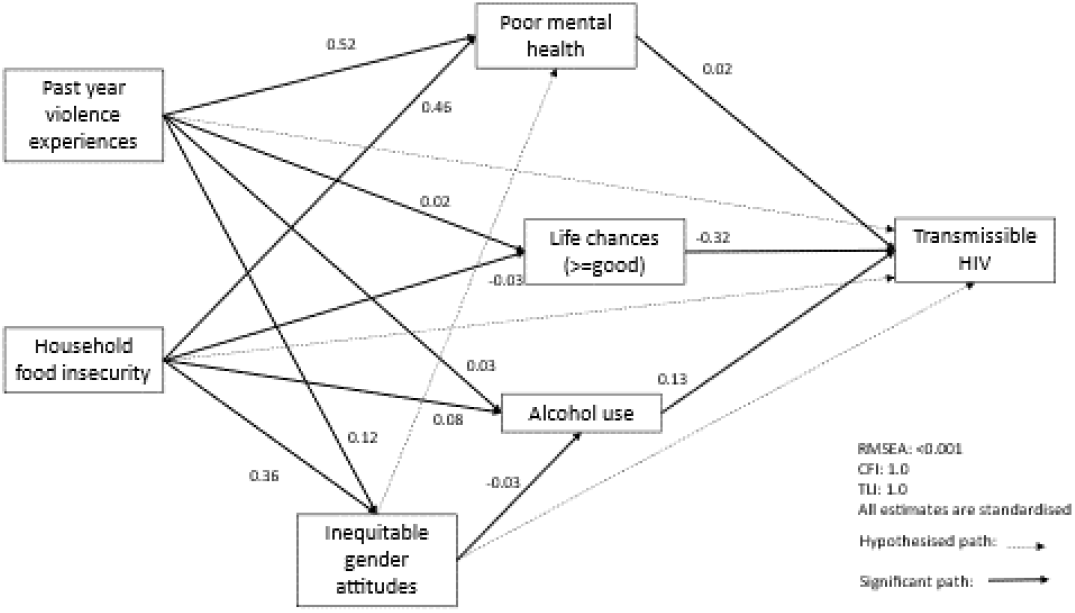
Path model for transmissible HIV and mediated pathways.

Men’s gender attitudes also mediated the relationship between food insecurity and violence experience, such that while violence experience and food insecurity were both associated with more gender inequitable attitudes, more gender inequitable attitudes were associated with greater alcohol use, which was in turn associated with transmissible HIV. Sensitivity models with different outcomes (HIV-negative status versus HIV-positive, HIV-negative status versus transmissible HIV) found the same relationships (Supplementary File: Table 5 and 6).

## Discussion

Among a population-based sample of young men (13-36 years old) living in rural South Africa one in twelve (8.5%) had sexually transmissible HIV. Notably, this comprised 78% of those living with HIV in the sample, suggesting the HIV treatment programme in its current format is ineffective for this group, reflecting previous research ^5^. Our analysis broadly supported our initial hypothesis that among young men, the likelihood of living with sexually transmissible HIV was shaped by recent violence experience and food insecurity, primarily mediated via poor mental health, as well as through the lack of a sense of life chances and alcohol use, although we were unable to adjust for age in the model.

Our analysis highlights the importance of the wider structural context of men’s lives in shaping their poor mental health and substance use, in turn impacting on transmissible HIV. Studies have often demonstrated poor mental health and substance use negatively impact on HIV treatment adherence ^18-20^, as well as HIV prevention ^38^. This has led to a focus on addressing young men’s poor mental health and substance misuse through individualised approaches, such as cognitive behavioural therapy or problem-solving approaches ^39,40^. While individualised treatments are important, our analysis highlights how poor mental health are a response to the wider social contexts of poverty and violence that young men live in. In 2018 in South Africa the unemployment rate for young men (15-34 years) was 38.2% ^41^. Moreover, young men experience exceedingly high rates of interpersonal violence, and are the group most likely to be murdered ^42^. Given our findings that these contextual factors indirectly impact on transmissible HIV, interventions to address HIV need to tackle these wider social factors as well as more proximate causes.

Our analysis also showed that perceptions of low life chances were strongly associated with transmissible HIV, and low life chances were shaped by greater food insecurity. Previous research has shown less hope is associated with poorer HIV treatment outcomes ^23^. It has been suggested that hope may be a way in which individuals make sense of, and interpret, the contextual constraints they face and something that interventions can directly address ^43^. In contrast to our hypothesis, men who experience more violence had higher perceptions of life chances. There are a variety of potential reasons for this. Research has shown hostility to people ‘doing better’ expressed via violence through to witchcraft ^44^, and it may be that men who were doing better in this setting, were more likely to experience violence. It could also be that men experience violence while being violent towards others – the use of violence by young men, particularly in settings of poverty, can be a way to assert dominance ^45,46^. As such it may be linked to a form of masculinity where success is partially indicated through the use of violence ^46^.

Rates of violence experienced by young men were very high in this population with a quarter reporting experiencing any form of violence and one in 35 men (2.8%) reporting experiencing sexual violence in the past 12 months and this did not vary by transmissible HIV status, but rather acted through other recognised pathways such as alcohol and poor mental health. Previous South African population-based data found 1 in 10 men reported lifetime sexual victimization by a man ^47^. There is strong evidence that experience of childhood sexual abuse is associated with worse health outcomes ^48^. Despite this relatively high prevalence, there remains little understanding around who perpetrators are and the underlying drivers of this sexual violence against men, suggesting this is an important area for future research, to enable a better understanding of the wider context of transmissible HIV in South Africa.

In the SEM there was a complex relationship between gender equitable attitudes and transmissible HIV. Food insecurity and experience of violence were associated with less equitable gender attitudes, reflecting prior research ^27^. There was however, a small but significant relationship between more gender inequitable attitudes and less alcohol use, in contrast to previous research which has suggested the opposite relationship ^49^. It is unclear why this relationship was seen, it may be that the limited measurement of ‘masculinity’ in the form of gender attitudes helps explain this. Qualitative research on the relationship between gender attitudes, masculinities and alcohol use in this context are required to understand this relationship.

This study has several limitations. Data were cross-sectional, with overlapping measurements for exposures and mediators. While we built models based on theoretical assumptions about the temporality of relationships, future research should assess the direction or directions of causality. We could not include age in the SEM, and age is a key confounder for many of these measures. We did not ask about who the perpetrators of violence were, nor the severity of violence experienced, and as such may be combining heterogenous experiences; developing more nuanced measures of men’s violence experience is important. Despite limitations, we used a random sample of men and a biological marker of transmissible HIV, enabling generalisation in this population. The setting is also somewhat typical of many rural parts of South Africa, and thus our findings may well generalize at least nationally.

This analysis highlights to reduce sexually transmissible HIV among young men in a rural context, we need to address the proximate ‘drivers’ of poor mental health and substance misuse, and also the structural drivers of these, namely food insecurity and experiences of violence, as well as how these manifest in perceptions of limited life chances. Building holistic interventions that adequately engage these multiple challenges is critical for improving HIV among young men.

## Data Availability

Data is available from: https://data.ahri.org/index.php/home

## Acknowledgements

We would like to acknowledge and thank the AHRI research team, including the research assistants, and research administrators for their effort and commitment to this study. We also would like to thank our research community, including the community advisory boards in uMkhanyakude district.

## Authorship

A.G., E.C. undertook the analysis. AG designed this analysis. N.C., J.D., C.H., N.O., O.B., N.M., K.B., T.Z., T.S, L.S., M.S. and J.S. designed the wider study this data drew on and oversaw data collection. A.G., M.S., E.C., G.H., L.S., N.M., J.S. contributed sections of the paper/wrote substantive components. N.C., J.D., C.H., N.O., O.B., N.M., K.B., T.Z., T.S. reviewed and edited the paper. All authors read and approved the final manuscript.

