## Supplementary Files for "eYoung men’s experiences of violence and poverty and the relationship to sexually transmissible HIV: a cross sectional study from rural South Africa"

Supplementary File

Supplementary Table 1: Measure of men’s experiences of violence. Responses yes/no.

**Has anyone ever done any of the following things to you in the last 12 months:**

**Emotional Violence Experience**

Say or do something to humiliate you in front of others?

Threaten to hurt or harm you or someone close to you?

Insult you or make you feel bad about yourself?

**Physical Violence Experience**

Push you, shake you, or throw something at you

Slap you

Twist your arm or pull your hair

Punch you with his fist or something that could hurt you

Kick you, drag you, or beat you up

Try to choke you or burn you on purpose

Threatened to attack you with a knife or other weapon

Attacked you with a weapon

**Sexual Violence Experience**

Touched you in a sexual way (e.g. kissing, grabbing, or fondling), when you did not want them to

Try to have sexual intercourse with you when you did not want to but did not succeed

Physically forced you to have sexual intercourse even when you did not want to

Forced you to perform sexual acts when you did not want to

Supplementary Table 2: descriptive comparison of those who consented to participate versus those who did not

|  | No | Yes |  |
| --- | --- | --- | --- |
|  | N(%) | N(%) | p-value |
| Consented into study | 1090(30.5) | 2487(69.5) |  |
| Age (mean, >=more) | 23.5(21.0, 26.0) | 21.2(21.0, 21.5) | 0.025 |
| Age (13/19) | 14(37.8) | 1287(51.8) | 0.146^a^ |
| 20/29 | 14(37.8) | 819(32.9) |  |
| 30/36 | 9(24.3) | 381(15.3) |  |
| ^a^ Fishers Exact test used | |  |  |

Supplementary Table 3: Descriptive comparison of those who provided dried blood spots (DBS) compared to those who did not.

|  | No | Yes |  |
| --- | --- | --- | --- |
|  | N(%) | N(%) | p-value |
| Provided HIV-data | 405(16.3) | 2082(83.7) |  |
| Age (Mean) | 23.9(23.2, 24.6) | 20.7(20.4, 20.9) | <0.001 |
| Education: Primary or less | 22(5.5) | 166(8.0) | <0.001 |
| Some secondary | 216(53.6) | 1447(69.6) |  |
| Secondary completed | 165(40.9) | 467(22.5) |  |
| Current situation: In school | 153(37.9) | 1257(60.5) | <0.001 |
| Out of school & unemployed | 145(35.9) | 488(23.5) |  |
| Out of school & part-time employed | 51(12.6) | 181(8.7) |  |
| Out of school & employed | 55(13.6) | 153(7.4) |  |
| Household food insecurity (>=more) | 0.43(0.34, 0.52) | 0.50(0.46, 0.54) | 0.18 |
| Poor mental health (>=more symptoms) | 3.49(3.12, 3.85) | 3.39(3.23, 3.55) | 0.633 |
| Poor mental health (yes) | 53(13.1) | 252(12.1) | 0.584 |
| Alcohol use: No use ever | 173(42.8) | 933(44.9) | 0.638 |
| Past month use | 137(33.9) | 657(31.6) |  |
| Binge drinking in past month | 94(23.3) | 490(23.6) |  |
| Life chances (>= more chance) | 2.56(2.51, 2.61) | 2.58(2.56, 2.60) | 0.408 |
| Gender attitudes (>=more inequitable) | 4.9(4.6, 5.2) | 5.6(5.5, 5.8) | <0.001 |
| Past year emotional violence experience (yes) | 50(12.4) | 390(18.7) | 0.002 |
| Past year physical violence experience (yes) | 43(10.6) | 385(18.5) | <0.001 |
| Past year violence experience (any - yes) | 69(17.0) | 561(27.0) | <0.001 |
| Any violence experience score (>=more) | 0.48(0.64, 0.79) | 0.71(0.64, 0.79) | 0.009 |

Table 4: Descriptive comparison between those HIV-negative, HIV-positive but undetectable viral load, and HIV-positive with transmissible viral load

|  | **HIV-negative** | **HIV-positive - undetectable viral load** | **HIV-positive - detectable viral load** |  |
| --- | --- | --- | --- | --- |
|  | n(%)/mean(95%CI) | n(%)/mean(95%CI) | n(%)/mean(95%CI) | p-value |
| **Socio-demographic factors** | 1853(89.0) | 51(2.5) | 178(8.6) |  |
| Age (Mean) | 19.9(19.7, 20.1) | 21.2(25.4, 29.0)** | 27.0(26.1, 27.9)** |  |
| Education: Primary or less | 149(8.1) | 4(7.8) | 13(7.3) | <0.001 |
| Some secondary | 1325(71.6) | 22(43.1) | 100(56.2) |  |
| Secondary completed | 377(20.4) | 25(49.0) | 65(36.5) |  |
| Current situation: In school | 1207(65.2) | 14(27.5) | 36(20.3) | <0.001 |
| Out of school & unemployed | 393(21.2) | 22(43.1) | 73(41.2) |  |
| Out of school & part-time employed | 139(7.5) | 9(17.7) | 33(18.6) |  |
| Out of school & employed | 112(6.1) | 6(11.8) | 35(19.8) |  |
| Household food insecurity (>=more) | 0.49(0.45-0.54) | 0.47(0.22, 0.72) | 0.55(0.39, 0.70) |  |
| **Mental health/substance use** |  |  |  |  |
| Poor mental health (>=more symptoms) | 3.30(3.14, 3.460 | 4.02(2.86, 5.12) | 4.15(3.56, 4.73)** |  |
| Poor mental health (yes) | 218(11.8) | 8(15.7) | 26(14.6) | 0.395 |
| Alcohol use: No use ever | 859(46.4) | 14(27.5) | 60(33.9) | <0.001 |
| Past month use | 580(31.3) | 20(39.2) | 57(32.2) |  |
| Binge drinking in past month | 413(22.3) | 17(33.3) | 60(33.9) |  |
| Life chances (>= more chance) | 2.60(2.58, 2.62) | 2.50(2.35, 2.64) | 2.43(2.36, 2.51)** |  |
| **Violence and attitudes** |  |  |  |  |
| Gender attitudes (>=more equitable) | 5.7(5.5, 5.8) | 4.5(3.5, 5.5)* | 5.3(4.8, 5.8) |  |
| Past year emotional violence experience (yes) | 345(18.6) | 11(21.6) | 34(19.1) | 0.86 |
| Past year physical violence experience (yes) | 347(18.7) | 6(11.8) | 32(18.0) | 0.443 |
| ^a^ Past year sexual violence experience (yes) |  |  |  |  |
| Past year violence experience (any - yes) | 497(26.8) | 13(25.5) | 51(28.70 | 0.847 |
| Any violence experience score (>=more) | 0.39(0.34, 0.44) | 0.33(0.02, 0.65) | 0.37(0.21, 0.52) |  |
|  | Continuous scored variables, compared to 'HIV-negative': *p<0.05 **p<0.01 | | | |
|  | ^a^ not compared by groups given small numbers in cells | | |  |

Table 5: Path model comparing those who are HIV-negative, with those living with HIV (no matter their viral load)

|  | **Estimate** | **SE** | **p-value** |
| --- | --- | --- | --- |
| Poor mental health -> Transmissible HIV | 0.02 | 0.01 | 0.021 |
| Life chances -> Transmissible HIV | -0.3 | 0.07 | <0.001 |
| Alcohol use -> Transmissible HIV | 0.16 | 0.05 | <0.001 |
| Household food insecurity -> Poor mental health | 0.46 | 0.07 | <0.001 |
| Violence experience -> Poor mental health | 0.52 | 0.04 | <0.001 |
| Household food insecurity -> Sense of life chances | -0.03 | 0.01 | <0.001 |
| Violence experience -> Sense of life chances | 0.02 | 0.01 | <0.001 |
| Violence experience -> Alcohol use | 0.03 | 0.01 | 0.001 |
| Household food insecurity -> Alcohol use | 0.08 | 0.02 | <0.001 |
| Inequitable gender attitudes -> Alcohol use | -0.03 | 0.01 | <0.001 |
| Violence experience -> Inequitable gender attitudes | 0.12 | 0.04 | 0.004 |
| Household food insecurity -> Inequitable gender attitudes | 0.36 | 0.08 | <0.001 |
| RMSEA =0.013; CFI=0.99, TLI=0.98 |  |  |  |

Table 6: Path model comparing those who are HIV-negative, with who have transmissible HIV

|  | **Estimate** | **SE** | **p-value** |
| --- | --- | --- | --- |
| Poor mental health -> Transmissible HIV | 0.02 | 0.01 | 0.022 |
| Life chances -> Transmissible HIV | -0.32 | 0.08 | <0.001 |
| Alcohol use -> Transmissible HIV | 0.14 | 0.05 | 0.005 |
| Household food insecurity -> Poor mental health | 0.45 | 0.08 | <0.001 |
| Violence experience -> Poor mental health | 0.52 | 0.05 | <0.001 |
| Household food insecurity -> Sense of life chances | -0.03 | 0.01 | 0.001 |
| Violence experience -> Sense of life chances | 0.02 | 0.01 | 0.014 |
| Violence experience -> Alcohol use | 0.03 | 0.01 | 0.003 |
| Household food insecurity -> Alcohol use | 0.08 | 0.02 | <0.001 |
| Inequitable gender attitudes -> Alcohol use | -0.03 | 0.01 | <0.001 |
| Violence experience -> Inequitable gender attitudes | 0.14 | 0.04 | 0.002 |
| Household food insecurity -> Inequitable gender attitudes | 0.36 | 0.08 | <0.001 |
| RMSEA =0.004; CFI=0.99, TLI=0.98 |  |  |  |
